# Interferon Gamma Release Assays for Latent *Mycobacterium tuberculosis* Detection in Elderly Hispanics

**DOI:** 10.1101/2021.06.15.21258953

**Authors:** Julia M. Scordo, Génesis P. Aguillón-Durán, Doris Ayala, Ana Paulina Quirino-Cerrillo, Eminé Rodríguez-Reyna, Mateo Joya-Ayala, Francisco Mora-Guzmán, Eder Ledezma-Campos, Alejandro Villafañez, Larry S. Schlesinger, Jordi B. Torrelles, Joanne Turner, Blanca I. Restrepo

## Abstract

**Background:** Aging is a tuberculosis co-morbidity. Interferon Gamma Release Assays (IGRAs) are used to detect latent *Mycobacterium tuberculosis* (*M*.*tb*) infection (LTBI) in adults, but their performance in the elderly is not well-established. We aim to evaluate the performance of IGRAs for LTBI detection in healthy elderly Hispanics with recent, remote or no history of *M*.*tb* exposure.

**Study Design and Methods:** Cross-sectional study in Hispanic elderly (60+y) and adult (18-50y) recent TB contacts (ReC) or community controls (CoC). LTBI was based on a positive T-SPOT.TB and/or QuantiFERON-Gold in-tube or –Plus assay.

**Results:** We enrolled 193 CoC (119 adults/74 elderly) and 459 ReC (361 adults/98 elderly). LTBI positivity increased with age in CoC (range 19-59%; trend p <0.001), but was similar in ReC (range 59-69%; trend p=0.329). The elderly had lower concordance between IGRAs (kappa 0.465 *vs*. 0.688 in adults) and more inconclusive results (indeterminate/borderline; 11.6% *vs*. 5.8% in adults; p=0.012). Exclusion of inconclusive results improved concordance between assays, notably in elderly ReC, who have the highest TB risk (from kappa 0.532 to 0.800). When both IGRAs were done simultaneously, inconclusive results were resolved in all cases as positive or negative with the other IGRA. The magnitude of the response to *M*.*tb* peptides used in the assays was similar between age groups, but responsiveness to mitogens was lower in the elderly.

**Conclusions:** IGRAs are suitable for LTBI detection in the elderly. Discordant and inconclusive findings are more prevalent in the elderly, but results were resolved when performing different IGRAs simultaneously in the same participant.

## INTRODUCTION

*Mycobacterium tuberculosis* (*M*.*tb*) infected nearly 10 million people and caused 1.8 million tuberculosis (TB) deaths worldwide in 2020.^1^ An estimated one quarter of the world’s population has a latent *Mycobacterium tuberculosis* (*M*.*tb*) infection (LTBI), with 5-10% anticipated to develop TB disease over their lifetime.^2^ The elderly have a 1.5-fold risk of progressing to TB when compared to adults, and are prone to severe treatment outcomes with 20-30% mortality.^3-5^ TB risk factors in the elderly include higher prevalence of LTBI and a dysfunctional immune system, marked by inflammaging and oxidative stress.^3,6^ Higher LTBI prevalence in older people is largely attributed to their lifetime-accumulated risk of exposure to *M*.*tb*.^7^

LTBI detection is based on immunological memory to *M*.*tb* peptides, measured using either the tuberculin skin test (TST) in-vivo or Interferon Gamma Release Assays (IGRAs) ex-vivo. The TST detects recall immunity to intradermal *M*.*tb* purified protein derivative antigens.^8^ IGRAs are based on IFN-γ production by peripheral blood mononuclear cells (PBMCs) in response to specific *M*.*tb* peptides.^9-11^

Interpretation of TST and IGRAs has been challenging in elders due to several factors. First, TST sensitivity is reported to decrease with older age ^12^, with higher false-negatives likely due to reduced skin immunity with aging.^13,14^ Second, LTBI is highly heterogeneous in the elderly, due to a range of recent or remote exposures to pulmonary TB patients. Third, studies comparing the performance of TST *vs*. IGRAs (T-SPOT.TB *vs*. QuantiFERONs) show conflicting results, with most reporting lower sensitivity of the TST, and among IGRAs, a higher sensitivity of the T-SPOT.TB *vs*. QuantiFERONs.^15-17^ When using active TB as a gold standard, most studies report lower sensitivity in the elderly with all assays, but a limitation of these studies is that most are done in hospitalized and severely ill patients.^16-19^ Another limitation of these studies is the use of different QuantiFERON kit versions that are no longer commercially available. Finally, most of these studies targeted elderly Asians, without data from Hispanics.

To assess the performance of IGRAs for LTBI detection in elderly Hispanics, we conducted a cross-sectional study comparing T-SPOT.TB and QuantiFERONs. We enrolled elderly community controls or contacts of recent TB patients, with similar groups of younger adults as reference. Our findings indicated a higher proportion of discordant, borderline or indeterminate findings in the elderly, but these inconclusive results were always resolved as positive or negative with the other IGRA. LTBI prevalence was similar with both IGRAs, and increased with age in the community controls, suggesting that IGRAs are suitable for the assessment of LTBI in the elderly. We provide further insight into the clinical interpretation of a positive IGRA in the elderly.

## METHODS

### Participant Enrollment

We enrolled adults (18-50 years) and elderly (>60 years) individuals in south Texas and northeastern Mexico between 2017 and 2021. These included: 1) Individuals with exposure to newly diagnosed TB patients for at least 5 hours within 6 months of enrollment (recent contacts; ReC); and 2) Community controls (CoC) with no reported exposure or a remote history of exposure (> 6 months prior to enrollment) to a TB case. In addition, we enrolled individuals between 51 to 59 years old, with the same enrollment and classification criteria as those mentioned for ReC or CoC. This study was approved by human subjects institutional review boards in Mexico (110/2018/CEI) and the United States (HSC-SPH-19-0308), and all participants signed a written informed consent.

### IGRAs

Blood was collected at the time of enrollment and simultaneously evaluated for LTBI with T-SPOT.TB (Oxford Immunotec) and a QuantiFERON test (Qiagen, Germantown). T-SPOT.TB evaluates the IFN-γ response of blood CD4 T cell lymphocytes to *M*.*tb* antigens ESAT-6 (panel A) or CFP-10 (Panel B), while the QFT-GIT evaluates these responses to the *M*.*tb* antigens ESAT-6, CFP-10, and TB7.7 (TB1).^11^ The QFT-Plus contains a second tube with *M*.*tb* peptides that stimulate CD8 T cells (TB2).^20^ During the course of this study Qiagen transitioned from the QuantiFERON Gold In-tube (QFT-GIT) to QFT-Plus (collectively referred to as QFT), with most participants tested with the QFT-Plus version (399/524, 76.1%). We did not find differences in their performance (**Supplementary Figure S1**). Indeterminate results with QFTs due to either low mitogen responses or high backgrounds were categorized according to the product inserts.^9^ When there was a borderline result with T-SPOT.TB or an indeterminate with QFT, the positive or negative result with the other IGRA determined the final LTBI diagnosis.

### Data Analysis

Data were analyzed using SAS vr. 9.4 (Cary, North Carolina). Chi-square or Fisher’s exact tests were used to compare categorical variables. Median values were compared by the Wilcoxon rank sum test. Comparisons of medians between more than two study groups were established by the Kruskall-Wallis test with post-hoc Dwass, Steel, Critchlow-Fligner (DSCF) method. The Cochran-Armitage trend test was used to assess significant changes over different age groups. Cohen’s kappa coefficient (k) was used to measure overall agreement between paired test results.^21^ P values were considered significant if ≤ 0.05, and borderline significant if <0.10. Graphs were plotted using GraphPad PRISM vr. 9.0.

## RESULTS

### IGRA prevalence across age groups

We enrolled 652 participants: 193 CoC (119 adults; 74 elderly) and 459 ReC (361 adults; 98 elderly), with sociodemographic and clinical characteristics similar to a subset of this cohort described recently (**Supplementary Table S1**).^22^ LTBI prevalence was significantly higher among elderly *vs*. adults in the CoC group (56.7% in elderly *vs*. 23.2% adults; p < 0.001), but similar between age groups of ReC (elderly 71.3% *vs*. adults 64.1%; p=0.233; **Table 1**). We also found few host characteristics associated with LTBI by univariable analysis (**Table 1**), with only macrovascular disease being more likely among the LTBI elderly in the ReC group (adj-OR 2.14, 95%CI 1.11, 4.12; **Supplementary Table S1**).^22^

**Table 1.** LTBI test results among elderly and young adults, by exposure history.

|  | Community & Recent Contacts |  |  | Community |  |  | Recent Contacts |  |  |
| --- | --- | --- | --- | --- | --- | --- | --- | --- | --- |
|  | Young adults | Elderly | P value | Young adults | Elderly | P value | Young adults | Elderly | P value |
|  | n (%) | n (%) |  | n (%) | n (%) |  | n (%) | n (%) |  |
| **T-SPOT.TB and QFT combined <sup>a</sup>** |  |  | **0.075** |  |  | **< 0.001** |  |  | 0.641 |
| Negative | 219 (45.6) | 65 (37.8) |  | 92 (77.3) | 33 (44.6) |  | 127 (35.2) | 32 (32.7) |  |
| Positive | 261 (54.4) | 107 (62.2) |  | 27 (22.7) | 41 (55.4) |  | 234 (64.8) | 66 (67.4) |  |
| **T-SPOT.TB** |  |  | 0.264 |  |  | **0.002** |  |  | 0.124 |
| Negative | 230 (48.0) | 72 (42.9) |  | 92 (77.3) | 37 (52.9) |  | 138 (38.3) | 35 (35.7) |  |
| Borderline | 19 (4.0) | 11 (6.6) |  | 2 (1.7) | 1 (1.4) |  | 17 (4.7) | 10 (10.2) |  |
| Positive | 230 (48.0) | 85 (50.6) |  | 25 (21.0) | 32 (45.7) |  | 205 (56.9) | 53 (54.1) |  |
| **QFT** |  |  | 0.117 |  |  | **< 0.001** |  |  | 0.177 |
| Negative | 173 (46.0) | 58 (39.5) |  | 58 (85.3) | 30 (53.6) |  | 115 (37.3) | 28 (30.8) |  |
| Indeterminate | 9 (2.4) | 8 (5.4) |  | 0 (0.0) | 2 (3.6) |  | 9 (2.9) | 6 (6.6) |  |
| Positive | 194 (51.6) | 81 (55.1) |  | 10 (14.7) | 24 (42.9) |  | 184 (59.7) | 57 (62.6) |  |
| **Borderline or indeterminate <sup>b</sup>** |  |  | **0.012** |  |  | 0.313 |  |  | **0.002** |
| Positive or negative | 452 (94.2) | 152 (88.4) |  | 117 (98.3) | 71 (96.0) |  | 335 (92.8) | 81 (82.7) |  |
| Borderline or indeterminate | 28 (5.8) | 20 (11.6) |  | 2 (1.7) | 3 (4.0) |  | 26 (7.2) | 17 (17.3) |  |

<sup>a</sup> Presence of LTBI was based on a positive T-SPOT.TB or QuantiFERON (QFT) assays (Gold In-tube or QuantiFERON-Plus).
<sup>b</sup> Borderline results with T-SPOT.TB or indeterminate with QFT. Differences between groups established by Chi-square test. Statistically significant or borderline significant differences are in bold text.

Next, we evaluated how LTBI prevalence changes with increasing age in CoC *vs*. ReC groups, by 10-year age intervals. For this analysis, we also included CoCs and ReCs who were between 51-59 years old (n= 111; **Supplemental material**). Results are shown for both IGRAs combined in **Fig. 1** (T-SPOT.TB and QFT) or for the T-SPOT.TB and QFTs alone in **Supplementary Figure S2**. IGRA positivity increased with older age among CoCs (trend p <0.001), but remained consistently higher than 60% throughout all the studied ages in the ReC group (trend p =0.329). Our results also indicated that the proportion of new *M*.*tb* infections among ReC was significantly higher in adults (mean 39.2%, 95%CI 29.6, 48.8%) when compared to the elderly (mean 11.4%, 95%CI 5.2, 17.6%; p <0.050; **Fig 1**). In summary, IGRA positivity increased with age in elderly CoCs, but remained consistently higher throughout all the age groups in the ReCs.

**Figure 1.**
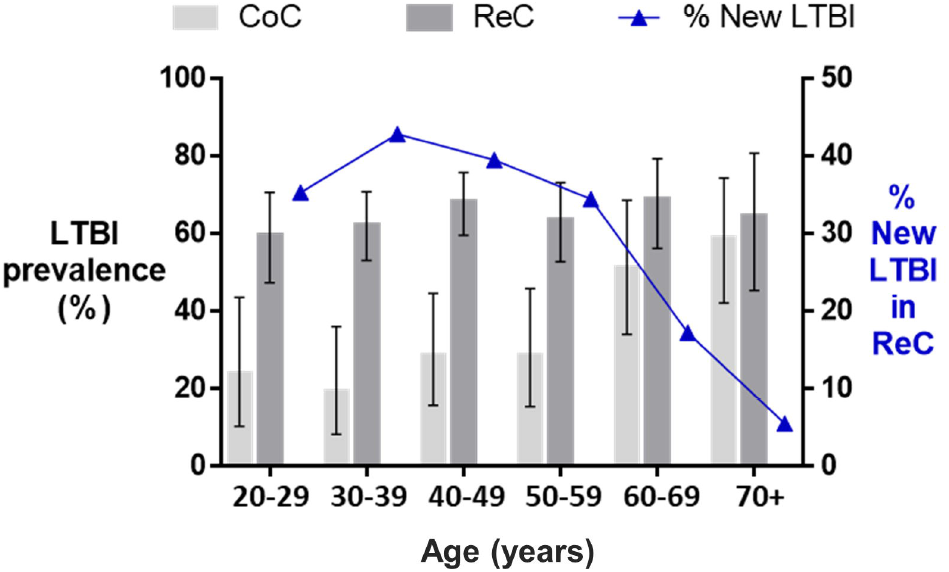
Changes in the prevalence of IGRA-positive results across age groups. The prevalence of LTBI was based on a positive T-SPOT.TB or QuantiFERON assay (QFT-GIT or QFT-Plus). The prevalence of LTBI increased with age among the CoC (*P* for trend = <0.001), but not for ReC (*P* for trend = 0.354). LTBI was significantly higher among CoC versus ReC up until 59 years old (non-overlapping 95% confidence intervals), but not among those 60 years or older. The proportion of newly-infected individuals in the ReC groups was calculated by subtracting LTBI-positives from the baseline CoC group (line with triangles). CoC, Community controls; ReC, Recent contacts; LTBI, latent *M. tuberculosis* infection. Vertical bars, 95% confidence intervals for LTBI prevalence.

### Concordance between IGRA assays

The proportion of elderly participants who tested IGRA positive was similar for T-SPOT.TB and QFT assays: 45.7% *vs*. 42.9% in the CoC group and 54.1% *vs*. 62.6% in the ReC group (**Table 1**). Among participants tested simultaneously with both types of IGRAs (518/652, 79.4%) in both CoC and ReC groups, the overall concordance between IGRAs was 80.4% (kappa 0.694; moderate; **Fig 2; Supplementary Table S2**). However, agreement between IGRAs differed by age groups. Among all elderly, the concordance was 71.4% (kappa 0.478; weak), *vs*. 83.8% in adults (kappa 0.695; moderate). When age groups were further stratified by CoC or ReC, IGRAs from the elderly also had a lower concordance when compared to adults (CoC kappa: 0.342 *vs*. 0.719 in adults; ReC kappa: 0.532 *vs*. 0.660 in adults; **Fig 2; Supplementary Table S2**). Agreement was lower for the CoC *vs*. ReC, and particularly low in the elderly (CoC: 65.4.2%, kappa 0.342, minimal; ReC: 74.8%, kappa 0.532, weak). In summary, discordant results were highest among the elderly, and particularly in the CoC group (at 34.6%, **Fig 2**).

**Figure 2.**
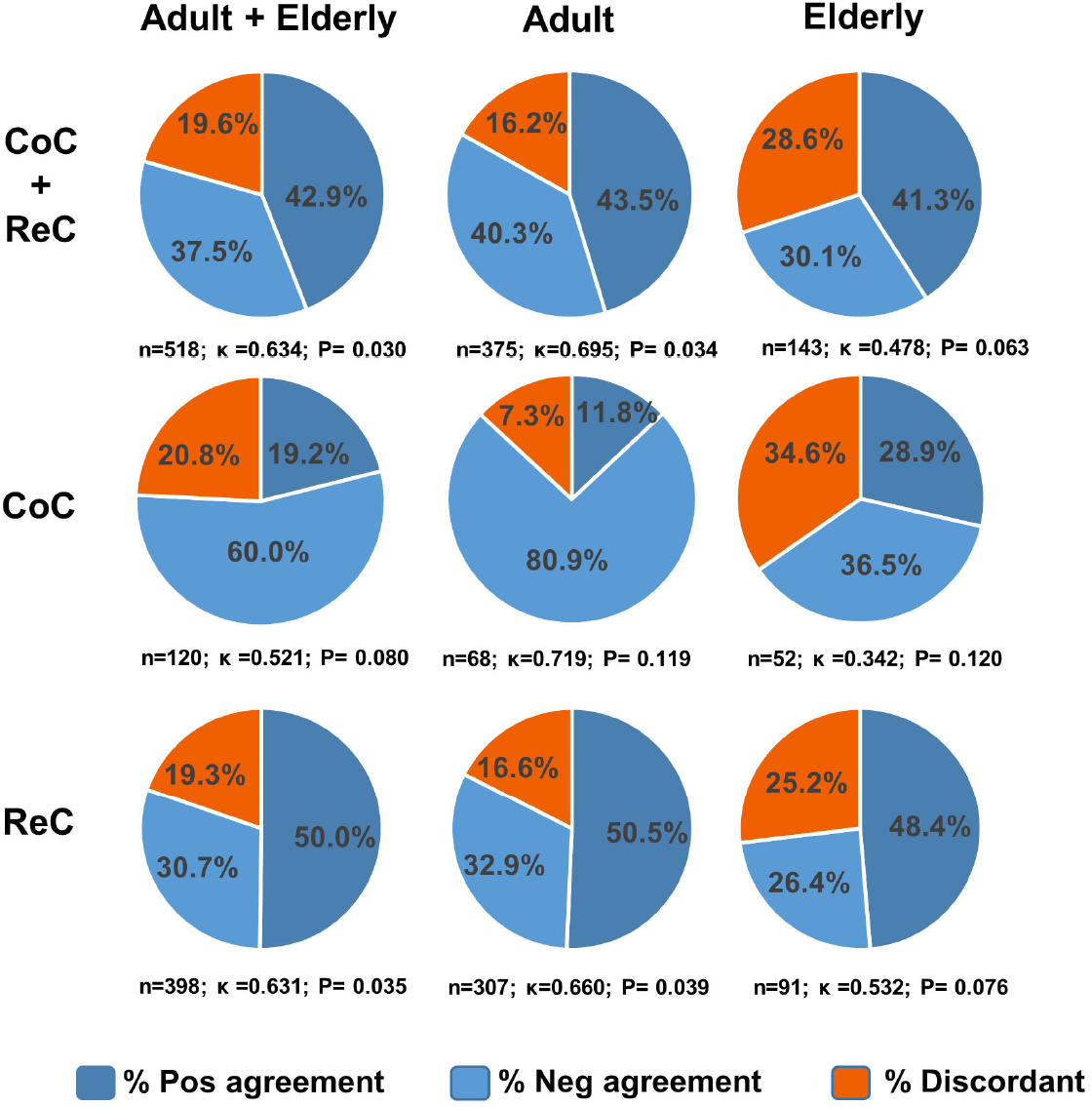
Concordance between IGRA assays, by age and study groups. The percent of positive and negative agreement was estimated for participants in whom both types of IGRA assays was done simultaneously (N=518/652, 79.4%), by age and study groups. The sample size and kappa statistic (k) is provided for each study group. CoC, Community controls; ReC, Recent contacts.

To determine the contribution of inconclusive results (indeterminate with QFTs or borderline with T-SPOT.TB) to the discordance between IGRAs, we excluded participants with inconclusive results and re-evaluated the data. The concordance between IGRAs improved in nearly all study groups (**Supplementary Table S2**). Among all participants, the agreement improved by 7.5% (from 80.4% to 87.9% after exclusions). The largest improvement was among elderly ReC / from 74.4% to 89.2%), the study group with the highest risk of TB.

### Interpretation of inconclusive results

Given the contribution of borderline or indeterminate results to the discordance between IGRAs, we expanded these analyses. Nearly all participants with borderline T-SPOT.TB results (n=28/30) and all with indeterminate QFT results (n=17/17) were tested with both IGRAs simultaneously. In all cases, the other IGRA resolved inconclusive findings, with all participants having a defined final classification as positive or negative LTBI (upper panel of **Table 1**). In comparison, if only one IGRA would have been performed per participant, the elderly would had higher proportion of borderline or indeterminate test results when compared to adults. This difference was significant among the ReCs group (elderly 17.3% *vs*. adults 7.2%; p=0.002), and had a similar trend for the smaller CoC group (elderly 4.0% *vs*. adults 1.7%; p=0.179; **Table 1)**. Thus, elderly individuals had a higher likelihood of an inconclusive LTBI result than adults.

We next determined if inconclusive results were more likely to be resolved as positive or negative with the complementary IGRA (**Fig 3A-B**). We found that a borderline T-SPOT.TB result was resolved as positive with the QFT in at least half the cases (14/28, 50% in adults; 7/11, 63.6% in elderly; **Fig 3**). In contrast, an indeterminate result by QFT was resolved as positive with the T-SPOT.TB in 100% of the cases: 17/17 among all participants tested, including 8/8 elderly. In all cases, indeterminate results were due to high IFN-γ background. In summary, borderline T-SPOT.TB results were true ‘borderline’, while indeterminate QFT results were always positive with the T-SPOT.TB.

**Figure 3.**
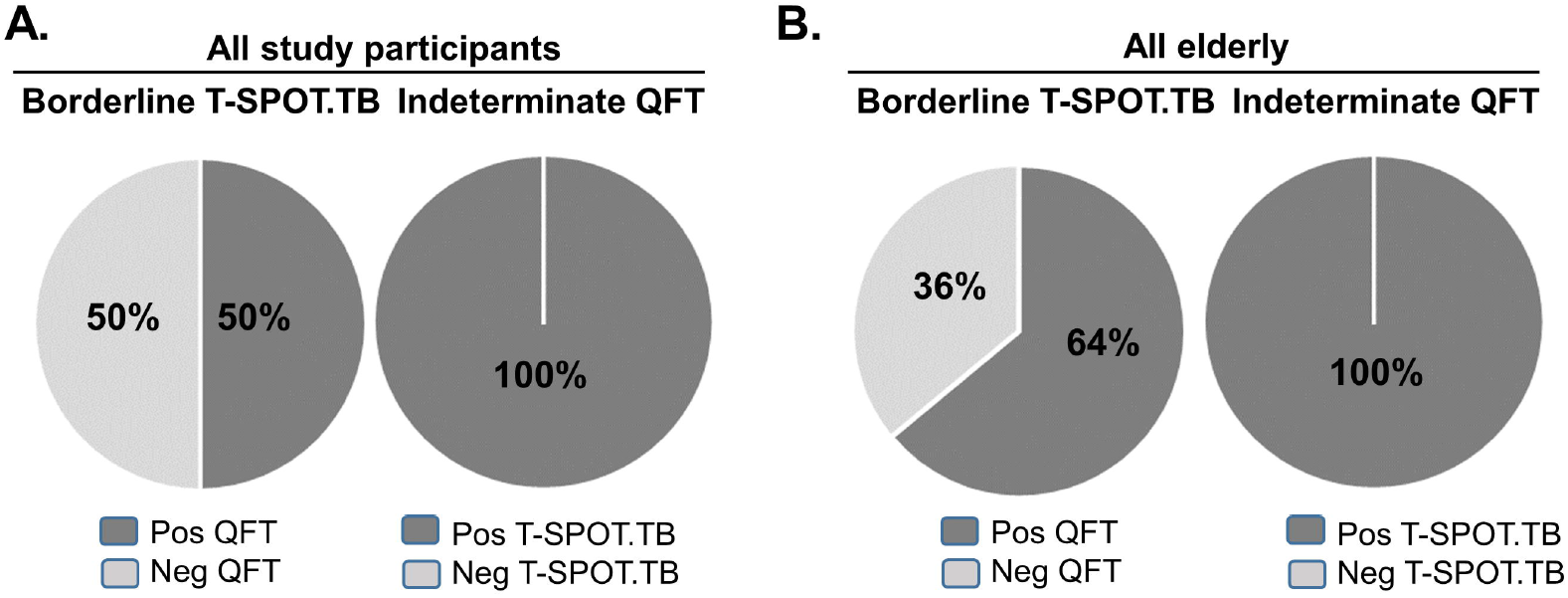
Resolution of inconclusive IGRAs. **A**. Among all study participants, borderline T-SPOT.TB was positive in 50% (14/28) of the cases by QFT, while intedeterminate QFTs were positive by T-SPOT.TB in 100% (17/17) of the cases. **B**. Among all elderly, borderline T-SPOT.TB was positive in 64% (7/11) of the cases by QFT, while intedeterminate QFTs were positive by T-SPOT.TB in 100% (8/8) of the cases.

### IFN-γ response among LTBI participants

We evaluated if there were differences in the magnitude of response to *M*.*tb* antigens (number of IFN-γ positive lymphocytes by T-SPOT.TB, or IFN-γ titers by QFT-Plus) by age group. Since the proportion of IGRA-positives was higher in the elderly, the analysis was limited to those positive with any assay. In the CoC group, there were no differences in the magnitude of IGRA responses to *M*.*tb* antigens between ages (**Table 2**). In the ReC group, the elderly had fewer lymphocytes responsive to the ESAT-6 antigen (p 0.05), but all other responses to *M*.*tb* antigens were similar between age groups. The only consistent alteration in the elderly *vs*. adults was their lower responsiveness to mitogens in the QFT-Plus assay, in both the CoCs (p 0.064) and ReCs (p 0.073). These findings were confirmed when expanding our analyses to all study participants, since the response to mitogens was not affected by the positivity of the IGRA (CoC p< 0.001; ReC p=0.035). Finally, we found that regardless of age, a recent exposure to a TB patient (ReC group) was associated with higher IFN-γ secretion *vs*. a remote exposure (CoC group; **Supplementary Table S3**). In summary, older age was associated with lower responses to mitogens, while the timing since exposure to an active TB case influenced the magnitude of the response to *M*.*tb* antigens.

**Table 2.** Magnitude of IGRA responses, among participants positive for LTBI ^a,b^.

|  | Community |  |  |  |  | Recent Contacts |  |  |  |  |
| --- | --- | --- | --- | --- | --- | --- | --- | --- | --- | --- |
|  | Adults |  | Elderly |  | P value | Adults |  | Elderly |  | P value |
|  | n | Median (IQR) | n | Median (IQR) |  | n | Median (IQR) | n | Median (IQR) |  |
| T-SPOT.TB |  |  |  |  |  |  |  |  |  |  |
| Nil | 29 | 0 (0.0) | 42 | 0 (1.0) | 0.281 | 244 | 0 (1.0) | 69 | 0 (0.0) | **0.023** |
| ESAT6 - Nil | 29 | 10 (21.0) | 42 | 10.5 (17) | 0.760 | 244 | 18 (19.0) | 69 | 10 (17.0) | **0.050** |
| CFP10 - Nil | 29 | 16 (18.0) | 42 | 20 (20) | 0.767 | 244 | 22 (17.0) | 69 | 22 (20.0) | 0.998 |
| QFT-Plus |  |  |  |  |  |  |  |  |  |  |
| Nil | 14 | 0.2 (0.6) | 31 | 0.2 (0.6) | 0.787 | 139 | 0.4 (1.4) | 44 | 0.5 (1.8) | 0.764 |
| Mit-Nil | 14 | 21.4 (16.0) | 31 | 15.4 (15.5) | **0.064** | 139 | 21.0 (11.6) | 44 | 15.5 (15.5) | **0.073** |
| TB1 - Nil | 14 | 0.4 (10.2) | 31 | 0.6 (1.8) | 0.806 | 139 | 4.2 (6.6) | 44 | 3.8 (6.5) | 0.436 |
| TB2 - Nil | 14 | 0.5 (8.2) | 31 | 0.7 (2.5) | 0.902 | 139 | 4.1 (7.1) | 44 | 3.9 (6.5) | 0.349 |
| Mit-Nil (all) <sup>c</sup> | 53 | 25.6 (13.9) | 56 | 15.5 (3.6) | <sup><</sup><br>**0.001** | 219 | 21.3 (11.5) | 71 | 16.1 (14.4) | **0.035** |

<sup>a</sup> Participants with positive IGRAs included those with borderline or indeterminate results who were classified as positive based on the other IGRA assay;
<sup>b</sup> Values from the indeterminate reactions were excluded due to their high non-specific IFN-γ background;
<sup>c</sup> Mitogen responses were also evaluated for all study participants, including those who tested IGRA-negative; Mit, mitogen; Group medians were compared by the Wilcoxon rank sum test; Statistically significant or borderline significant differences are in bold text.

## DISCUSSION

We evaluated the performance of IGRAs among Hispanic elderly *vs*. adults in ReC and CoC groups. This design allowed us to compare the performance of the T-SPOT.TB and QFT assays across age groups, and between participants with recent *vs*. remote exposure to active TB cases. Our design contrasts with most previous studies, where IGRAs were evaluated among active TB cases across age groups, or as part of outbreak investigations among the elderly.^15-17^ Furthermore, our analysis was done with QFT-Plus, in contrast to previous studies that evaluated earlier versions of QFT.^16,17,23^

There is a concern that the decline in the strength of the immune response in the elderly may decrease the sensitivity of assays to detect LTBI in this population.^24^ This limitation is observed for the TST ^12,17^, with a higher proportion of false negatives attributed to compromised skin immunity in the elderly.^13^ A lower sensitivity has also been reported for IGRAs in the elderly, although less marked than with the TST. However, these observations are from studies conducted amongst active TB patients, and with false-negatives associated with very advanced TB disease, lymphopenia or malnutrition.^16,17,25^ Evaluation of IGRAs in the elderly with TB in these studies may differ from results we obtained in those without TB for several reasons. First, our elderly participants did not have severe underlying conditions that could compromise their immunity, other than a high prevalence of diabetes that was not associated with their LTBI status (**Supplementary Table S1**). Second, previous studies in participants with TB also reported higher indeterminate results in the elderly, but these were due to low mitogen responses ^19,25^, while our indeterminate results were due to high IFN-γ background. Third, we observed an increasing prevalence of LTBI positives with age, even among individuals older than 60 years (**Fig 1**). However, we did not have a gold standard for TB infection since active TB patients were not studied, and hence, cannot rule out a lower sensitivity in the oldest participants (70 years+). We think this is unlikely because we did not detect a notable defect in the magnitude of the cell-mediated immune responses to *M*.*tb* antigens in the elderly when compared to adults (**Table 2**), and the prevalence of LTBI was similar when using T-SPOT.TB *vs*. QFTs (**Supplementary Figure S2C**). Together, our results suggest that the sensitivity of IGRAs for the detection of LTBI cases is not significantly impacted by old age alone.

Previous studies report challenges with IGRAs in the elderly *vs*. adults, namely poorer concordance between IGRAs, or between IGRAs and the TST.^15-17^ Studies have also reported a higher proportion of indeterminate results in older people.^17,25-27^ In our cohort, we also found that in the elderly *vs*. adults, there was a lower concordance between IGRAs, and a higher proportion of indeterminate or borderline results. We found a relationship between both observations, with discordant results being largely attributed to inconclusive results. These unclear results hinder a physician’s decision to recommend LTBI treatment, in particular to elderly ReC who have a high risk of progression to active TB disease, and in whom inconclusive results were the highest (17% of ReC elderly; **Table 1**). Importantly, in our cohort we found that the resolution of inconclusive results was possible in 100% of cases with a complementary IGRA, yielding a positive or negative result in a timely manner. The resolution of inconclusive results led to improved concordance in all study groups, including in the elderly ReC: from a kappa of 0.532 (weak) to 0.800 (strong; **Supplementary Table S2**). Thus, resolution of inconclusive results by testing with a second IGRA is a time-efficient alternative when compared to the routine retesting of ReCs 8-10 weeks later to confirm if there was a true conversion to a positive LTBI result.^28^

The higher risk of active TB development among the elderly has been largely attributed to their waning immune response.^24^ For decades, this hypothesis has been based in part on data showing the lower responsiveness of the elderly to the TST. Our results using IGRAs do not provide support for a marked immunocompromised response to *M*.*tb* antigens in the elderly. First, we confirmed that the prevalence of LTBI increases with age in the community, when using IGRAs that evaluate cell-mediated immunity to *M*.*tb*. Second, the magnitude of the response to *M*.*tb* antigens in the CoC or ReC group was comparable between the elderly and adults who were LTBI positive, and is consistent with a previous report.^25^ Third, the higher proportion of indeterminate results with the QFT in the elderly was due to a high IFN-γ background in all cases, and not to low mitogen responses. Thus, higher indeterminate results in the elderly may be a reflection of their baseline inflammatory status (inflammaging), or immune hyper-responsiveness upon exposure to *M*.*tb* antigens.^6^ The lack of an evident compromise in T-cell responses to *M*.*tb* in the elderly is consistent with findings from a recent study by our group.^29^ Nevertheless, the lower responsiveness of the elderly to the T-cell mitogen in the QFT-Plus assay (**Table 2**) suggests a generalized defect in cell-mediated immunity, which is consistent with their reduced responsiveness to vaccines.^30^

The higher prevalence of LTBI in the elderly is thought to be a major contributing factor to their increased risk of active TB development.^3,31^ However, in our cohort, the LTBI prevalence flattened at around 70%, with a significantly lower proportion of new infections among the elderly *vs*. the adults (**Fig 1**). Thus, we posit that elderly individuals who are not markedly immunocompromised, may have a higher resistance to LTBI conversion or active TB disease development compared to adults. There are several observations supporting this possibility. First, a recent historical review of the literature among individuals of all ages confirmed that active TB is more likely to develop shortly after initial infection, with a dwindling risk up to two years, and rarely thereafter. Even though a case for the elderly is not explicitly made, a bimodal distribution for the risk of TB reactivation is not observed over time.^32^ Second, there are studies suggesting that LTBI-positive individuals may in fact be more protected from re-infection with *M*.*tb*.^7,33^ Third, the interpretation of a positive LTBI result in individuals with remote exposure is unclear, with some having a robust recall response to a past LTBI but no longer harboring viable *M*.*tb*.^32^ Fourth, the elderly are more likely to have been exposed to *M*.*tb* during their lifetime, even multiple times, and hence, are a select groups of survivors of active TB and LTBI.^7^ Thus, the higher prevalence of positive LTBI tests among the elderly as a group is not necessarily a risk factor for active TB disease development, except for individuals who still harbor dormant bacilli in granulomas and are immunocompromised. Further studies are needed to understand the relationship between a positive LTBI test result and the risk of progression to active TB disease, particularly for elderly contacts from decades ago. These studies are critical to guide decisions on LTBI treatment in the elderly population.^14,34^

We recognize study limitations. First, there is no gold standard for LTBI assessment, so we based LTBI on positivity with any of the two IGRAs tested. Second, we switched from the QFT-GIT to the QFT-PLUS kit version during the course of our study, although 76.1% of the participants were evaluated with the – PLUS kit and the performance of both kit versions was similar (**Supplementary Figure S1**). The inclusion of both kits, therefore, allowed for additional analyses in our cohort; and in our hands, the additional TB2 tube in the QFT-PLUS had a minor improvement in the sensitivity (about 2%; **Supplementary Table S4**). This is expected, since the TB2 tube was mostly added to improve sensitivity in TB patients.^35^ Third, the cross-sectional nature of our design did not allow for the identification of the subset of elderly participants with recent IGRA conversion, who are thought to have the highest risk of progression to active TB disease. Finally, our study included elders with comorbidities such as diabetes and macro- and micro-vascular diseases, but data cannot be extrapolated to those severely immunocompromised.

Overall, our results suggest that IGRAs are not affected by waning immunity in the elderly, which contrasts with previous studies using the TST, or studies in the elderly with active TB. Furthermore, despite a higher proportion of discordant or inconclusive results in the elderly *vs*. adults, the resolution of inconclusive results was always possible when using a complementary IGRA. Thus, IGRAs are suitable to evaluate LTBI in the elderly; however, despite the high prevalence of LTBI in the elderly, the interpretation of a positive LTBI test (TST, IGRAs) should be interpreted with caution prior to consideration of LTBI treatment.^14^

## Data Availability

All data is available.

## Abbreviations

IGRAs: Interferon Gamma Release Assays
LTBI: latent *Mycobacterium tuberculosis* infection
TB: tuberculosis
ReC: recent TB contacts
CoC: community controls
QFT: QuantiFERON
TST: tuberculin skin test

## Acknowledgements

We would like to acknowledge Kristen Maynard, Danyelle Garza, Erica de Leon, Marielena Benavidez, Jose A. Caso, Juan Carlos Lopez-Alvarenga and Fabiola Lopez for technical support. We also thank health professionals and administrators at the Secretaría de Salud de Tamaulipas, including Q.F.B. Cristela Resendez-Cardoso, Drs. Francisco Garcia-Luna Martinez and Ariel Mercado-Cárdenas (administration) and Mr. Jorge Perez-Navarro (logistics). We also thank health professionals at the Casa Club del Adulto Mayor, Sistema DIF Matamoros. We thank the study participants for their time and interest in this study. Finally, we dedicate this study to the memory of our team members who were passionate about TB research, and whom we lost to COVID-19 in 2020, Dr. Francisco Mora-Guzmán and R. Eminé Rodriguez-Reyna.

## Author contributions

**Julia M. Scordo:** Original draft, data analysis, review and editing; **Genesis P. Aguillón-Duran, Eminé Rodríguez-Reyna, Alejandro Villafañez**: Data collection and curation; **Doris Ayala, Ana Paulina Quirino-Cerrillo, Mateo Ayala-Joya**: Data curation and specimen processing; **Larry Schlesinger, Jordi B. Torrelles**: Conceptualization, review & editing; **Joanne Turner**: Conceptualization, Project administration, Resources, review & editing; **Eder Ledezma-Campos, Francisco Mora-Guzmán**: Project administration, review & editing; **Blanca I. Restrepo** (guarantor): Conceptualization, Project administration, data analysis, review & editing. **All authors**: Reviewed and approved the final version.

