## Supplemental Information for "Interferon Gamma Release Assays for Latent *Mycobacterium tuberculosis* Detection in Elderly Hispanics"

**Supplemental material**

**Methods**

**Differences between the QFT-GIT and QFT-Plus versions**

The differences between the QFT-GIT and QFT-Plus kit versions have been described previously [1]. Namely, the QFT-GIT evaluates CD4 T cell responses to a mixture of *M.tb* peptides from the ESAT6, CFP10 and TB7.7 antigens (TB1 tube). In the QFT-Plus the TB1 tube had the TB7.7 antigen removed since it did not contribute to improved sensitivity, and an additional tube, TB2, was added. TB2 contains similar peptides to the TB1, plus additional peptides that stimulate CD8 T cells. This addition was aimed at increasing the sensitivity of this assay for active TB case detection [1].

**Comparison of the QFT-GIT vs. QFT-Plus**

Results were similar between both kit versions of QFT in adults (QFT-GIT n=104; QFT-Plus n=272) or elderly (QFT-GIT n=20; QFT-Plus n=127; **Supplemental** **Figure S1**). Given these similar findings, data from participants evaluated with both QFT kit versions were included in this study.

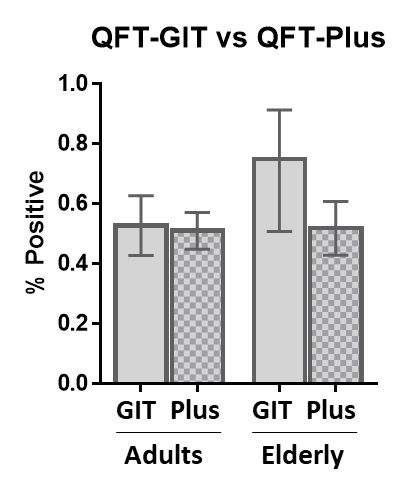
 **Supplemental** **Figure S1. Similar results with the QFT-GIT (GIT) and QFT-Plus (Plus) among adults or elderly.** Bars indicate mean proportions (% positive) with 95% CI.

**Results**

**Description of the study population**

Characteristics of the 652 participants (480 adults and 172 elderly) are shown in **Supplemental Table S1**. 67.5% were females, only 40.8% had an education beyond middle school, and only 27.8% were current or past smokers. Two-thirds were obese or overweight (75.3%), 23.2% had macrovascular disease and 25.0% had diabetes (98% type 2 diabetes). Most were BCG vaccinated (89.2%).

For the analysis by 10 year interval age groups we also included individuals 51-59 years old (n=111) enrolled during the same period of time as the other study participants. They comprised mostly females (71.2%), 43 had diabetes (38.7%) and 55.0% were LTBI-positive.

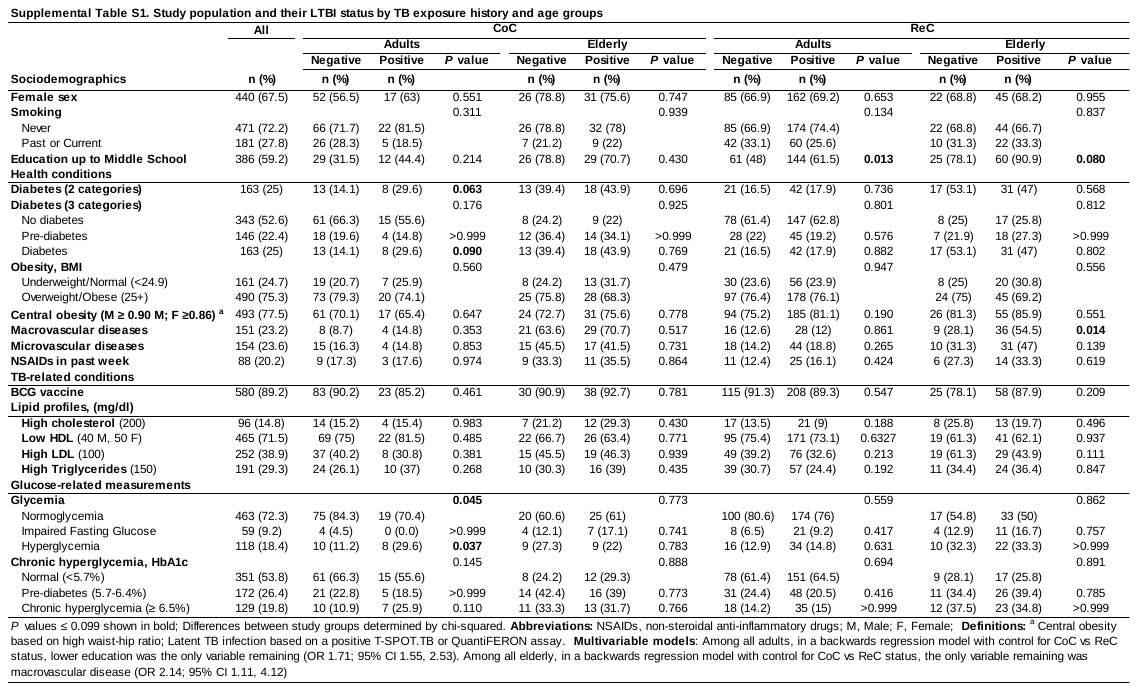

**Prevalence of by 10y age intervals, using the T-SPOT.TB or QFTs (Supplemental Figure S2)**

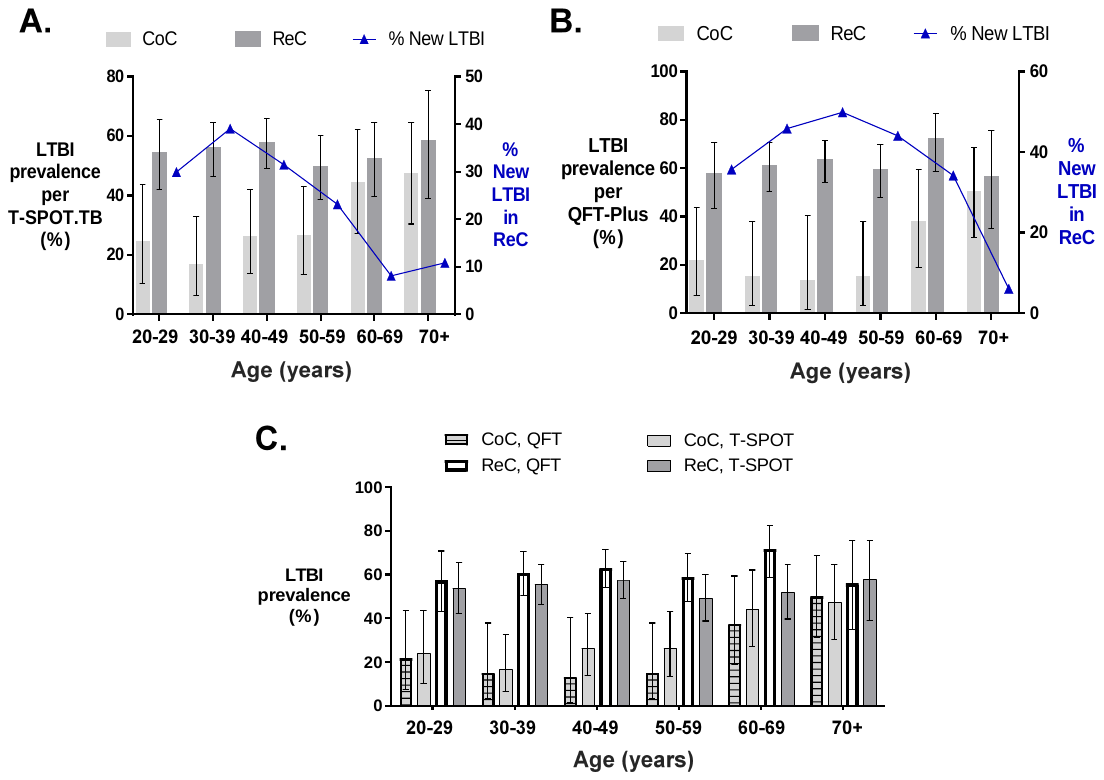

**Supplemental Figure S2. Individual and comparative analysis of LTBI prevalence with the T-SPOT.TB and QFT assays.**

**A-B.** The prevalence of LTBI was based on a positive T-SPOT.TB or QuantiFERON assay (QFT-GIT or QFT-Plus). The prevalence of LTBI increased with age among the CoC (*P* for trend: T.SPOT-TB p = 0.003; QFT *P* = <0.001), but not for ReC (*P* for trend: T.SPOT-TB *P* = 0.435; QFT p= 0.438). LTBI was significantly higher among CoC versus ReC in most of the younger age groups (non-overlapping 95% confidence intervals), but not among those 60 years or older. The proportion of newly-infected individuals in the ReC groups was calculated by subtracting LTBI-positives from the baseline CoC group (line with triangles). Vertical bars, 95% confidence intervals for LTBI prevalence. **C.** Similar prevalence data for LTBI using QFTs (QFT-GIT or QFT-Plus) or the T.SPOT-TB, shown side by side for each age groups. Overlapping confidence intervals indicate there is not a significant difference in the results obtained with both types of assays. CoC, Community controls; ReC, Recent contacts; LTBI, latent *M. tuberculosis* infection. Vertical bars, 95% confidence intervals for LTBI prevalence.

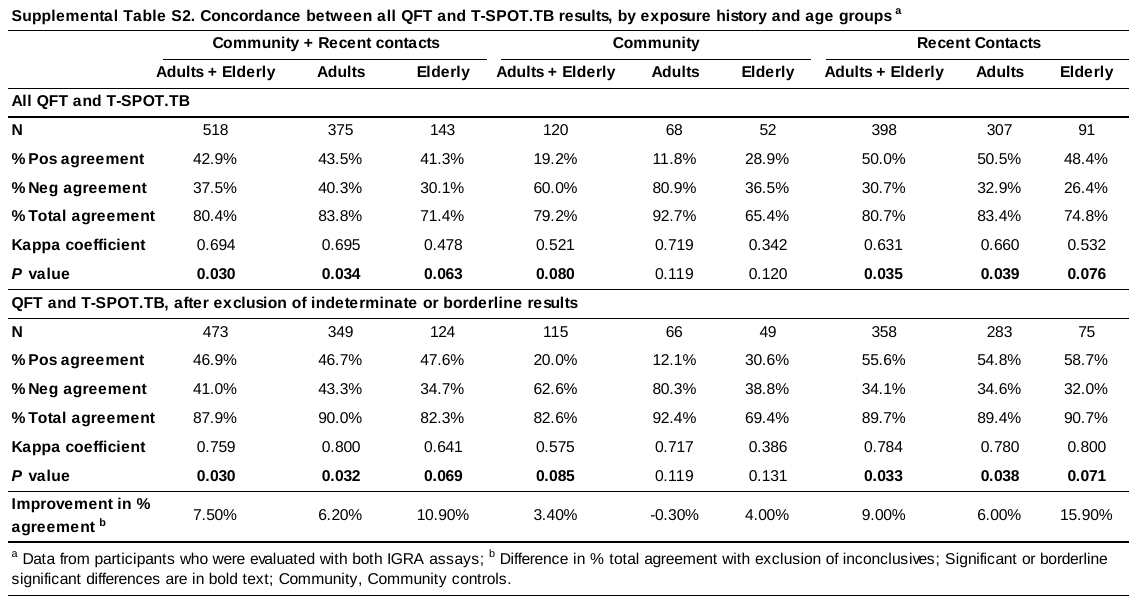

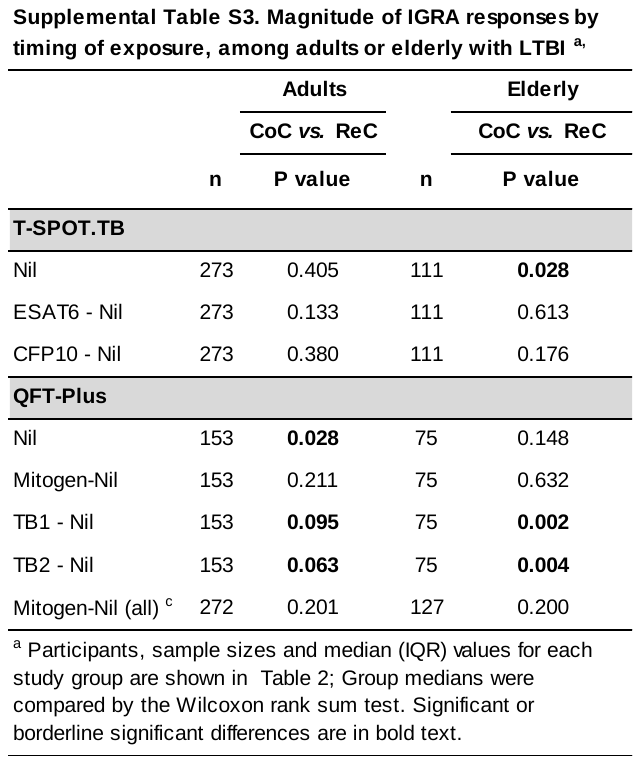
